# Redefining Macrosomia Risk Factors in Low-Resource Settings: A Cross-Sectional Study from an Ethiopian Tertiary Hospital

**DOI:** 10.1101/2025.08.05.25332915

**Authors:** Hailu Chare Koyra, Ashebir Tesfaye Tufa, Eskindir Wolka Woticha, Atnafu Nega Nadew

**Affiliations:** School of Pharmacy, Wolaita Sodo University, P.O. Box 138, Wolaita Sodo, Ethiopia; School of Medicine, Wolaita Sodo University, P.O. Box 138, Wolaita Sodo, Ethiopia; School of Public Health, Wolaita Sodo University, P.O. Box 138, Wolaita Sodo, Ethiopia; School of Nursing, Wolaita Sodo University, P.O. Box 138, Wolaita Sodo, Ethiopia

**Keywords:** Wolaita Sodo, macrosomia, risk factors, South Ethiopia, contraceptive use

## Abstract

**Background:** Fetal macrosomia (birth weight ≥4000g), as defined by WHO, poses significant risks for maternal and neonatal health, particularly in low-resource settings. In Ethiopia, where obstetric services face substantial challenges, the predictors of macrosomia extend beyond traditional risk factors like diabetes and obesity. This cross-sectional study at a tertiary hospital in South Ethiopia aimed to investigate underrecognized determinants of macrosomia, to inform targeted antenatal risk stratification. The findings address a critical evidence gap in the region and provide actionable insights for clinical practice.

**Methods:** We conducted a facility-based cross-sectional study at Wolaita Sodo University Comprehensive Specialized Hospital (WSUCSH) from October 1 to December 30, 2020, involving 467 randomly selected postpartum women. Data were collected through structured questionnaires and medical record review, capturing maternal characteristics and birth outcomes. After data entry using EpiData version 3.1, we performed statistical analysis in SPSS 25, including: descriptive statistics for baseline characteristics, bivariate logistic regression to identify potential predictors, and multivariate logistic regression to determine independent associations with macrosomia. Statistical significance was set at p<0.05 with 95% confidence intervals.

**Result:** The study revealed a macrosomia prevalence of 15.8% among 467 deliveries. Three key predictors emerged: male fetal sex (AOR=2.71, 95%CI:1.22-5.98), prior macrosomic delivery (AOR=8.53, 95%CI:2.64-27.62), and hormonal contraceptive use (AOR=3.57, 95%CI:1.30-9.79). Notably, injectable contraceptives accounted for 54% of family planning methods among mothers of macrosomic infants.

**Conclusion and recommendation:** This study identifies hormonal contraception as a novel, modifiable risk factor for macrosomia in low-resource settings, alongside established predictors (male sex, prior macrosomia). The findings advocate for: (1) enhanced antenatal screening for contraceptive history, (2) targeted fetal monitoring for high-risk pregnancies, and (3) provider education on non-traditional macrosomia risks. These evidence-based interventions could significantly reduce adverse outcomes in resource-limited hospitals.

## 1. INTRODUCTION

The World Health Organization (WHO) defines macrosomia as absolute birth weight of 4000 grams and above; regardless of gestational age. In developed countries, the most commonly used threshold is weight above 4500 g. There is grading system, which may be useful at term for decision-making regarding operative delivery has been suggested: grade 1 for infants 4000 to 4499 g, grade 2 for 4500 to 4999 g, and grade 3 for over 5000 g [9].

Uncontrolled diabetes mellitus in pregnancy, maternal obesity, excessive gestational weight gains as well as maternal over nutrition have been associated with fetal macrosomia. Other risk factors of macrosomia include male fetal sex, high parity, maternal height and post-term pregnancy [15,16,17].

Locally we define fetal macrosomia as birth weight equal to or greater than 4000 grams [31]. Due to these varying definitions and variety of their contributory factors, it has a wide prevalence of 0.5% to 20% of all pregnancies worldwide [31, 34]. In Ethiopia study done at Hawassa city, south Ethiopia indicate the prevalence of fetal macrosomia 11.86%, retrospective cross-sectional study done in Mekele city indicate the prevalence of 19.1% [32,34].

Fetal macrosomia is a major contributor to obstetric morbidity. Due to the maternal and neonatal morbidities associated with pregnancies of macrosomic fetuses, such pregnancies are often termed high-risk pregnancies [2]. Fetal macrosomia, which is found in all parts of the world, is expected to be higher in affluent countries where their nutritional levels are higher [2]. The prevalence of fetal macrosomia in the developed countries has specifically increased by 15-20% recently; an increase largely attributed to increasing maternal obesity and diabetes [4].

Macrosomic infants are prone to still births, birth asphyxia, meconium aspiration syndrome, skeletal injuries, brachial plexus injury, hypoglycemia, hypocalcaemia, clavicle fracture, respiratory distress, Polycythemia, increased frequency of NICU admission, low Apgar score increase and shoulder dystocia. It also causes long term complications like obesity, impaired glucose tolerance and metabolic syndrome during childhood and beyond [9, 29].

Maternal complications associated with macrosomic delivery include obstructed labor, uterine rupture, perineal injuries, post-partum haemorrhage, prolonged labor, instrument-assisted delivery, Caesarean sections and prolonged hospital stay [29].

Fetal macrosomia poses a significant obstetric challenge because antenatal diagnosis can be problematic and inaccurate. Increased index of suspicion based on knowledge of predictors of fetal macrosomia may assist in selecting women, in which antenatal fetal weight estimation is important. [29,33]. Macrosomia can be a greater obstetric hazard for women in developing countries like Ethiopia where under nutrition during childhood can inhibit growth of the pelvis to its full potential, pregnancy before the pelvis is fully developed is common, and facilities for operative delivery of women with obstructed labor are not consistently available.

There is a need for increased documentation of the associated factors and outcome of pregnancies complicated by fetal macrosomia in the study area, thus necessitating this study. In particular, magnitude and associated factors of fetal macrosomia are not well examined in Wolaita Sodo University teaching Referral Hospital. In addition, there is no published study done at Wolaita, South Ethiopia. Meanwhile, a better understanding of associated factors of fetal macrosomia in this particular setting is essential for designing specific cost-effective interventions aimed at reducing macrosomia complications. Therefore, the main aim of this study was to assess magnitude and associated factors of fetal macrosomia among mothers delivered at Wolaita Sodo University teaching referral hospital, South Ethiopia, 2020.

Despite this burden, there is no published study done at Wolaita, South Ethiopia to assess the magnitude and associated factors of fetal macrosomia among mothers delivered which is a basic strategy to prevent fetomaternal complication associated with fetal macrosomia. Previous studies done at different regions of the country shows figures with wide discrepancies regarding the factors associated with fetal macrosomia. In addition, in most of the studies conducted in our country, the associated factors of fetal macrosomia assessed by retrospective cross-sectional method, where under reporting or missing charts is possible. This research used prospective cross sectional method of study, which potentially avoids the aforementioned gaps. In addition previous studies done in Ethiopia cannot evaluate the association between maternal chronic medical problem, substance abuse and contraceptive use with fetal macrosomia. Because the availability of local information on the associated factors has major role to control this severe public health problem, this study assessed magnitude and associated factors of fetal macrosomia in WSUTRH. Therefore, the result of this study will help policy makers, program planners, governmental and non-governmental organization implementers and maternal health service providers/practitioners to provide evidence-based interventions, which will reduce the burden of fetomaternal complication of fetal macrosomia.

Most importantly, since there are limited research evidences on the magnitude and associated factors of fetal macrosomia in the study area despite the high burden of associated complication, this study will serve as a baseline work for other researchers interested to work on further researches in this topic.

## 2. METHODS AND MATERIALS

### Study design, area and period

The study was conducted from November 1 to December 30, 2020 GC at WSUTRH in Wolaita zone, which is found 396 km south from Addis Ababa and 165km far from Hawassa, south Ethiopia. The Hospital specifically found in southern nation nationalities people’s government Region, Wolaita zone, Sodo Town. Its catchment population is five million, from which male account 49% and female 51%. WSUTRH was established in 1920 E.C and zonal hospital in Wolaita zone, Sodo town till June 30/2004 E.C after which time incorporated to Wolaita

Sodo university and currently serving as teaching and research centre in addition to its curative and rehabilitative care with in the hospital as outpatient, and in patient, as well as at community level in outreach services. The hospital has one big maternity ward, which possesses around 60 beds. There are about 6000 deliveries per year, 30 % of which is caesarean deliveries. Some of the services which are given by these departments are pre-operative and post-operative, in patient services, abortion care and safe abortion services, labor and delivery services, PMTCT services, ART services for all pregnant women and Obstetric/Gynaecologic Ultrasound services. There are 7 obstetricians, 36 residents and 25 midwives currently working in the department of obstetrics [35]. An institution based cross sectional study was conducted at WSUTRH from November 01-December 30, 2020 G.C.

### Population

all labouring mothers who delivered at WSUCSH from November 01-December 30, 2020 G.C were source population and the study participants were women gave singleton deliveries irrespective of gestational age, at WSUTRH.

### Inclusion criteria

Women who gave birth (live or still birth) with birth weight of greater than 1000gram

### Exclusion criteria

Participants who unable to respond because of medical/obstetric complications, woman less than 18 years that cannot give consent, woman with multiple pregnancies and other who delivered a baby (live or stillbirth) with severe visible congenital anomaly

### Sample size determination

The required sample size was determined using single population proportion formula [30] with the assumption by taking the magnitude of fetal macrosomia 19.1% [32], 95% level of confidence and 3% marginal error. n is sample size, Z (α/2)^2^ is critical value with Z α/2 = 1.96, d is precision = 0.03 (margin of error), P =19.1 %(0.191) since the magnitude of fetal macrosomia at Mekele university was 19.1% [32].

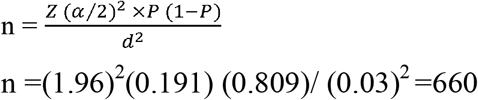

One month total delivery is about 600, which becomes 1200 in two months delivery. Since the source population for this study is less than 10,000 adjustment is made nf= n/ (1+n/N), 660/ (1+660/1200) =424.5, by adding 10% contingency the sample size was 424.5+ 42.45= 467 samples.

### Sampling procedure

The study participants were selected through simple random sampling technique. Accordingly, women who delivered at WSURTH during the data collection period were consecutively included in the study until the desired sample size was achieved based on the eligibility criteria.

### Data collection procedure

Data was collected by two first year residents and supervised by one senior resident. The data was collected using structured questionnaire. Information regarding sociodemographic character, obstetric history and pregnancy related factors were extracted both from participants and their documents. Meanwhile the completeness of the data was checked every day by the principal investigator and supervisor. Principal investigator was made regular supervision and follow up for completeness and consistency of the data on daily basis. After identification of the participants, they were approached and explained for the purpose of the study and were given consent to participate.

### Data processing and analysis

Data was coded and entered into Epidata version 3.1 and then exported in to SPSS 25 for analysis, both bivariate and multivariate analysis was computed. The p value of less than 0.05 was considered as the level of significance. The result was described using tables, pie charts and other graphs.

### Data quality control

before collecting data, the checklist was tested to check the consistency of the checklist format and the data collector’s performance. The checklist was modified based on the pretest results. Orientation on how to carry out data collection was given for the data collectors. One day training was given to data collectors about the objective of the study, methods of data collection and how to collect important information. Pretest was conducted at Dubo St. marry general hospital on the 23 mothers (5 %) of the sample size. Accordingly, the consistency, understandability and completeness of the questionnaires was insured based on pre-test result. The principal investigator was in control of the overall data collection process. Data was entered in Epi data computer programs to minimize data entry error.

### Ethical clearance

Before data collection, ethical clearance and approval was obtained from institutional review Board (IRB) of the Wolaita University, college of health science and medicine. Communication with the medical director and department head was made through formal letter obtained from school of medicine and permission was obtained from medical director and department head to cascade the research. After the purpose and objective of the study was informed, verbal and written consent was obtained from each study participants. In order to keep confidentiality of any information provided by study subjects, the data collection procedure was anonymous. Participation was on voluntary basis and they can withdraw from the study at any time. To ensure the confidentiality of the respondents their names was not written on the questionnaires.

## 3. RESULTS

### 3.1. Maternal socio demographic characteristics

A total of 467 eligible mothers who delivered at WSUTRH from 1^st^ November 2020 to 30^th^ December 2020 were asked to participate in the interview. All of the mothers gave full information, the response rate of 100%. From total of 467 respondents about half of them 243 (52%) were urban. Almost all 463 (99.14%) were married and 425 (91.0%) were between the ages of 20–34 years. The mean age of mothers was 27.32 ±3.985. Two-thirds 357(76.44%) of respondents were Wolaita in ethnicity. More than half, 237(51.28%) had primary education (table 1).

**Table 1:** Socio-demographic characteristics of mothers delivered at WSUCSH from November 1 to December 30, 2020(N=467).

| S.N | Variables(N=467) |  | Frequency | Percentage |
| --- | --- | --- | --- | --- |
| 1. | Age | Less than 20 | 32 | 6.9 |
|  |  | 20-34 | 425 | 91.0 |
|  |  | Greater than 35 | 10 | 2.1 |
| 2. | Marital status | Single | 2 | 0.43 |
|  |  | Married | 463 | 99.14 |
|  |  | Divorced | 2 | 0.43 |
| 3. | Religion | Orthodox | 142 | 30.40 |
|  |  | Muslim | 31 | 6.64 |
|  |  | Protestant | 273 | 58.46 |
|  |  | Other | 21 | 4.50 |
| 4. | Ethnicity | Wolaita | 357 | 76.44 |
|  |  | Gamo | 22 | 4.71 |
|  |  | Goffa | 19 | 4.07 |
|  |  | Hadiya | 19 | 4.07 |
|  |  | Other | 50 | 10.71 |
| 5. | Educational status | Illiterate | 105 | 22.50 |
|  |  | Primary school | 237 | 51.28 |
|  |  | High school | 57 | 12.20 |
|  |  | College & above | 68 | 14.56 |
| 6. | Occupation | Housewife | 235 | 50.32 |
|  |  | Merchant | 71 | 15.20 |
|  |  | Private employee | 63 | 13.50 |
|  |  | Government employee | 46 | 9.85 |
|  |  | Student | 32 | 6.85 |
|  |  | Other | 20 | 4.28 |
| 7. | Monthly income of respondents | Less than 1000birr | 83 | 17.8 |
|  |  | 1001-2000birr | 96 | 20.5 |
|  |  | 2001-4000birr | 113 | 24.2 |
|  |  | Greater than 4000birr | 175 | 37.5 |

**Table 2:** Pregnancy related factors among mothers delivered at WSUCSH from November 1 to December 30, 2020(N=467)

| S.N | Variables (N=467) |  | Frequency | Percentage |
| --- | --- | --- | --- | --- |
| 1. | ANC follow up | Yes | 445 | 95.30 |
|  |  | No | 22 | 4.70 |
| 2. | Where was ANC follow up | Health center | 234 | 52.58 |
|  |  | Primary hospital | 27 | 6.07 |
|  |  | Private institution | 121 | 27.19 |
|  |  | WSUCSH | 63 | 14.16 |
| 3. | Number of ANC visit | Once | 5 | 1.12 |
|  |  | Two times | 29 | 6.52 |
|  |  | Three times | 76 | 17.08 |
|  |  | Four and above | 335 | 75.28 |
| 4. | Parity of the respondents | One | 150 | 32.12 |
|  |  | Two to four | 243 | 52.03 |
|  |  | More than or equal to 5 | 74 | 15.85 |
| 5. | Screening for diabetes mellitus | Yes | 13 | 2.78 |
|  |  | No | 454 | 97.22 |
| 6. | Birth out come | Alive | 461 | 98.7 |
|  |  | Stillbirth | 6 | 1.3 |
| 7. | Weight gain during pregnancy | Less than or equal to 5kg | 1 | 0.2 |
|  |  | 6-10kg | 16 | 3.4 |
|  |  | Greater than 11kg | 180 | 38.6 |
|  |  | No weight measurement | 270 | 57.8 |

### 3.2. Pregnancy related factors

Majority 445(95.3%) of respondents had ANC follow up, only 22(4.7%) mothers were not attended ANC follow up. About half 234(52.58%) of mothers were attended their ANC follow up at health center and three-fourth 335(75.28%) of mothers visited four or more than four times. About one-third 150 (32.12%) of mothers were primiparus and 317(67.88%) of mothers were multi-parus. Regarding the gestational age, only 2(0.43%) of neonates were born pre-terms, 139(29.76%) of neonates were born at terms, 41(8.78%) of neonates were born post-term and majority 285(61.03%) gestational age was unknown (Table 3 and figure 2).

Only 13 (2.8%) of mother were screened for gestational diabetes, 454 (97.2%) of mothers were not screened. Among screened mother 3 (23%) of them were diabetic and 10 (77%) of them were non-diabetic. Regarding maternal weight gain only 1 (0.2%) mothers had 5kg and less weight increment, 16 (3.4%) of mothers had 6 to 10 kg weight increment, 180 (38.5%) of them had weight increment between 11to 15 kg and more than half 270 (57.8%) had no weight measurement at all (table2).

### 6.3. Fetal macrosomia and risk factors

In this study, 74(15.85%) of delivered fetus were macrosomic and about four-fifth, 392 (83.94%) of the new born baby had a normal birth weight (2.5–3.9 kg) (figure 1). More than half, 270 (57.81%) of the women had no preconception Body Mass Index (BMI) or first trimester weight. Most, 461 (98,7%) of neonates were alive and more than half, 241 (51.6%) of neonates were male. All most all 454(97.22) of the respondents were not diagnosed with any chronic disease. Only one-sixth 74 (15.85) of respondents were grand multipara. Moreover, 423 (90.58%) of the women had no history of macrosomic baby (Table 3).

**Figure 1.**
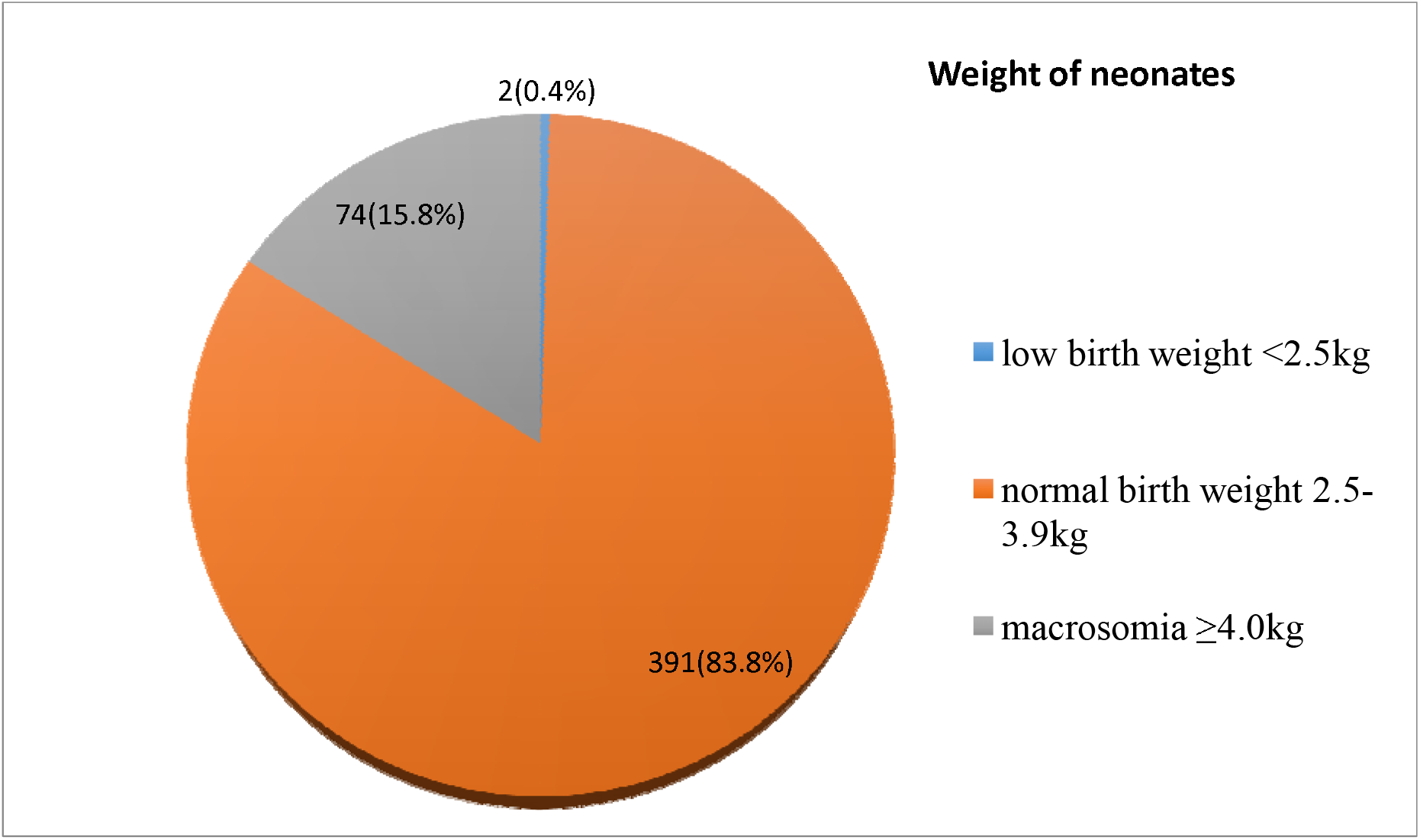
Weight of neonates delivered at WSUCSH from November 1 to December 30, 2020 (N=467).

**Figure 2.**
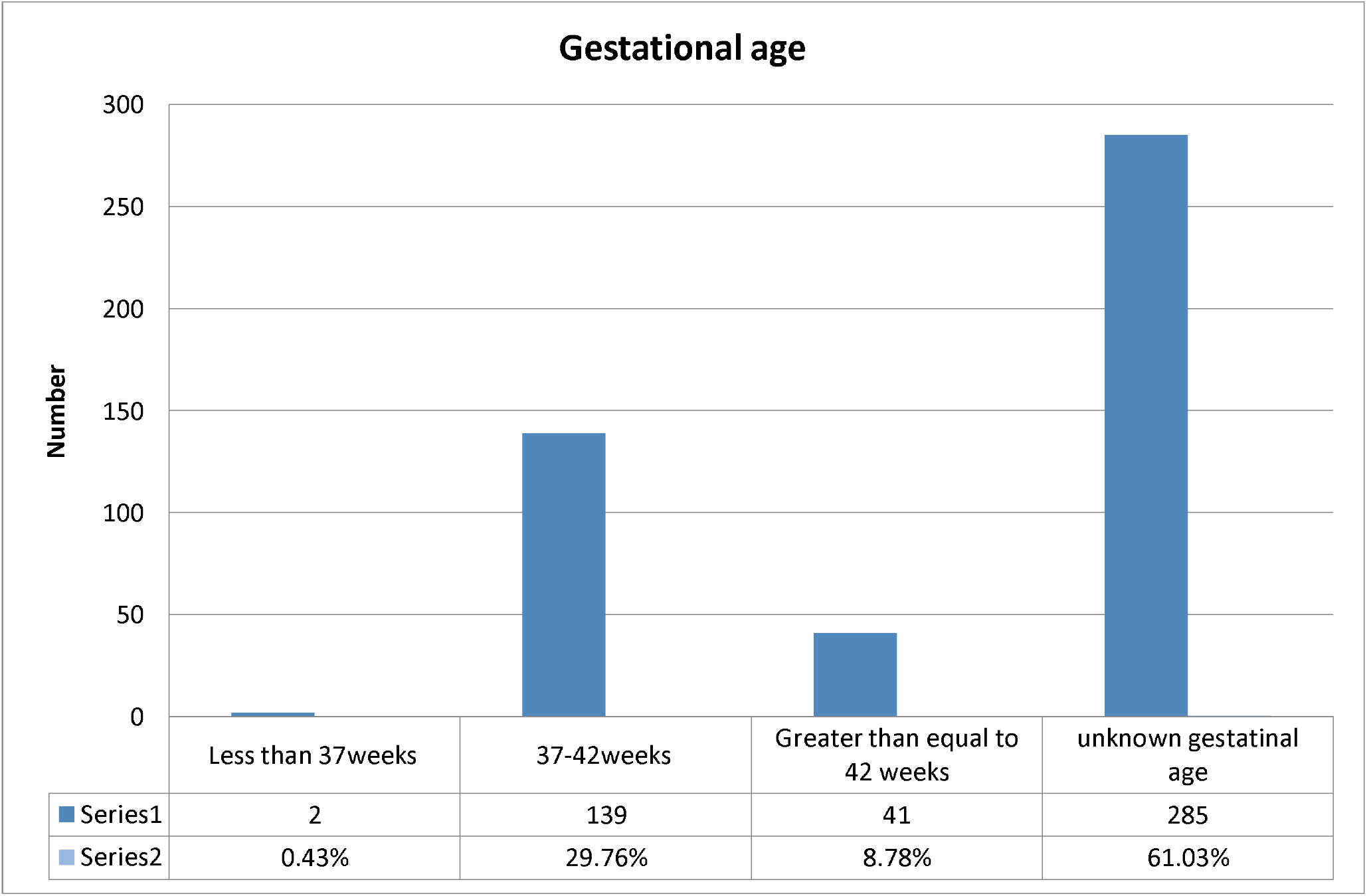
Gestational age during delivery among mothers delivered at WSUTRH from November 1 to December 30, 2020(N=467).

### Factors associated with fetal macrosomia

In the bivariate analysis the factors found to be significantly associated with fetal macrosomia were: Educational status, Occupation, ANC follow up, Gestational age at time of delivery, sex of the child, gestational age, previous history of fetal macrosomia, screening for DM, current weight of mother, history of family planning and monthly income. However, in the multiple logistic regression analysis: sex of the child, previous history of fetal macrosomia and history of family planning were significantly associated with the magnitude of fetal macrosomia.

Being a male was about 2.7 times more likely to be associated with fetal macrosomia than being a female (AOR = 2.705, 95% CI 1.224-5.980). Similarly, having previous history of fetal macrosomia was about 8.5 times more likely to be associated with fetal macrosomia than no macrosomia history (AOR = 8.534, 95% CI 2.637-27.624). Moreover, having history of family planning also significantly associated with fetal macrosomia (AOR = 3.573, 95% CI 1.304-9.788) (Table 4).

**Table 4:** Factors associated with fetal macrosomia among mothers delivered at WSUCSH from November 1 to December 30, 2020(N=467)

| S.N | Variables(N=467) |  | Frequency | Percentage |
| --- | --- | --- | --- | --- |
| 1. | History of DM | Yes | 3 | 0.64 |
|  |  | No | 464 | 99.36 |
| 2. | Sex of neonate | Male | 241 | 51.6 |
|  |  | Female | 226 | 48.4 |
| 3. | Chronic medical problem | Yes | 13 | 2.78 |
|  |  | No | 454 | 97.22 |
| 4. | Type of medical problem (N=13) | Diabetes mellitus | 3 | 23.08 |
|  |  | Hypertension | 5 | 38.46 |
|  |  | HIV | 5 | 38.46 |
| 5. | Period between previous pregnancy and latest | No previous pregnancy | 144 | 30.83 |
|  |  | Less than 1 year | 71 | 15.20 |
|  |  | 1 to 2 years | 142 | 30.41 |
|  |  | More than 2 years | 110 | 23.56 |
| 6. | History of family planning | Yes | 272 | 58.2 |
|  |  | No | 195 | 41.8 |
| 7. | Type of family planning used by respondents | Pills | 16 | 5.88 |
|  |  | Injection | 147 | 54.04 |
|  |  | Implants | 76 | 27.94 |
|  |  | IUCD | 8 | 2.94 |
|  |  | Others | 25 | 9.20 |
| 8. | Previous history of fetal macrosomia | Yes | 44 | 9.42 |
|  |  | No | 423 | 90.58 |

**Table 5:** Comparison of associated factors with fetal macrosomia among mothers delivered at WSUCSH from November 1 to December 30, 2020(N=467)

| Variables<br>(N=467) | Category | Fetal weight |  | COR | AOR | P-value |
| --- | --- | --- | --- | --- | --- | --- |
|  |  | Macrosomic | Non- macrosomic |  |  |  |
| Sex of neonate | Male | 52 | 189 | 2.551(1.492-4.362) * | 2.705(1.224-5.980) * * | 0.014 |
|  | Female | 22 | 204 | 1.00 | 1.00 |  |
| Previous history of fetal macrosomia | Yes | 26 | 18 | 11.285(5.764-22.095) * | 8.534(2.637-27.624) * * | 0.000 |
|  | No | 48 | 375 | 1.00 | 1.00 |  |
| Current weight of mother | Less than 70kg | 32 | 283 | 1.00 | 1.00 |  |
|  | ≥70kg | 42 | 110 | 0.296(0.178-0.493) * | 1.360(0.513-3.607) | 0.536 |
| Gestational age at time of delivery | Less than 37 weeks | 0 | 2 | 0.000 | 1.00 |  |
|  | 37-42 weeks | 22 | 117 | 1.119(0.637-1.965) | 0.777(0.291-2.071) | 0.613 |
|  | ≥42 weeks | 11 | 30 | 2.182(1.014-4.694)* | 0.308(0.089-1.067) | 0.063 |
|  | Unknown | 41 | 244 | 1.00 | 1.00 |  |
| History of family planning | Yes | 51 | 221 | 1.726(1.015-2.935) * | 3.573(1.304-9.788) * * | 0.013 |
|  | No | 23 | 172 | 1.00 | 1.00 |  |
| *COR, Statistically significant **AOR= statistically significant, p<0.05. |  |  |  |  |  |  |

## 4. DISCUSSION

This study attempted to determine the magnitude and associated factors of fetal macrosomia at Wolaita Sodo University Comprehensive Specialized hospital, Southern Ethiopia.

The magnitude of fetal macrosomia was found to be 15.8%. This finding is within global range, which is 0.5% to 20% [1, 2,3]. This study is consistent with the studies conducted in Indian range of prevalence, 7.7% in normal-weight women and 12.7% in obese women [18], Algeria (14.9%) [23], Uganda (8.4%) [22], and southern Ethiopia Hawassa city (11.8%) [34], moreover, the present study finding is lower than the studies done in Mexico (18.6%) [14] and Mekelle city, Tigray region of Ethiopia (19.1%) [32]; while, it is higher than the studies conducted in 14 provinces in China (7.3%) [5], United States of America (8.07%)[8], Ghana 6.5%[23], Nigeria (5.5%) [3] and Tanzania (2.3 %) [20]. This variation might be due to difference in the socio-demographic characteristics of the mothers, study design, data collection procedure and health care difference of the facilities. For instances, the study done in Ghana was conducted in rural community that may decrease prevalence of fetal macrosomia due to low socioeconomic status [23]. Similarly, study done in Mekele city and Tigray region of Ethiopia was conducted in the private clinics where mothers with better socioeconomic status get the service, which could increase the magnitude of macrosomia [32].

In the current study, there is a significant association between sex of the child and fetal macrosomia. Those children who were male had 2.7 times greater odds of macrosomia than those who were female. This is similar with studies conducted in Japan [10] and Cameroon [17]. Moreover, previous history of macrosomia was significantly associated with fetal macrosomia. This finding is in line with the studies conducted in Iran [13] and Hawassa city of Ethiopia [34].

This study showed that using family planning was 3.5 times more likely associated with fetal macrosomia. Even if there were no studies showed that the positive relationship between history of family planning and fetal macrosomia, there are studies that shows indirect relationship of family planning use and obesity. There are many studies, which showed direct relationship of obesity and fetal macrosomia [16, 19, 25, 26]. Study done in United States of America has also shown that Depo-Provera users gain on an average 4.4 kg in 2 years and 5.1 kg in 3 years. In addition, Depo-Provera users also have an increase in visceral fat, which is most metabolically active in promoting dyslipidaemia. These findings suggest that Depo-Provera recipients should be monitored for BMI after 6 month and regularly thereafter [21].

Even this is a plausible scientific assumption for association between family planning and fetal macrosomia, for which further studies need to create true relationship with appropriate design. The finding of this study revealed that only 2.78% of mothers were screened for gestational diabetes and only 197 (42%) mothers were having weight measurement. More than half 270 (57.8%) had no weight measurement at all, hence it was difficult to calculate BMI and weight gain during pregnancy for the rest participants. But most studies showed gestational diabetes and maternal weight gain during pregnancy were strong predictors of macrosomia [15, 16]. This study fail to test for this associated factors due to failure of mothers to recall their pre-pregnancy or first trimester weight, documentation gap or weighing mothers and screening mothers for diabetes is less practiced in the institutions. Even though there was difficulty to relate BMI with fetal macrosomia, it showed that there was strong association of BMI with fetal macrosomia among those with pre-pregnancy weight measurement.

Additionally, most of mothers delivered at study facility didn’t know gestational age during delivery, which made it impossible to show the association with fetal macrosomia. There were many literatures showed that relationship of gestational age during delivery with fetal macrosomia [9, 12, 20].

Unlike other studies, factors like, maternal age, parity, occupation, monthly income, chronic medical problem, maternal height, maternal weight at time of delivery, substance abuse and nutritional history were not significantly associated with neonatal birth weight in this study.

### Conclusion and Recommendations

This study identified three significant predictors of fetal macrosomia: male fetal sex, previous history of macrosomia, and hormonal contraceptive use (particularly injectable methods). These findings highlight the need for enhanced antenatal risk assessment protocols in low-resource settings. We recommend: (1) implementation of routine gestational diabetes screening and maternal weight monitoring across all antenatal clinics; (2) establishment of referral networks that ensure complete transfer of maternal health data, including pre-pregnancy weight and gestational age; and (3) targeted fetal growth monitoring for women with prior macrosomic deliveries or using hormonal contraception. Healthcare providers should emphasize early and regular antenatal care to facilitate timely risk identification. While this study revealed important associations, further research is needed to: (a) investigate the causal mechanisms between hormonal contraceptives and fetal overgrowth, and (b) evaluate the immediate maternal and neonatal complications of macrosomia in this setting through longitudinal study designs. These evidence-based interventions and research priorities will enable better prevention and management of macrosomia-related adverse outcomes.

### Strength and Limitations of the study

This prospective cross-sectional study provides novel insights into hormonal contraceptive use as a predictor of macrosomia, addressing a critical evidence gap in South Ethiopia. Its design minimized recall bias and documentation gaps through direct data collection. However, several limitations warrant consideration: (1) unavailable gestational age data and pre-pregnancy weight measurements precluded analysis of these established risk factors; (2) the cross-sectional design cannot establish causal relationships; and (3) findings from this tertiary hospital may not generalize to community settings. These limitations highlight the need for longitudinal studies incorporating comprehensive antenatal measurements.

## Disclosure

Authors declare that they have no competing interests for financial or other conflict of interest in this work

## Data Availability

All data produced in the present study are available upon reasonable request to the authors

## ABBREVIATIONS AND ACRONYMS

ACOG: [American College of Obstetricians and Gynecologists]
[BMI]: Body Mass Index
[GDM]: Gestational Diabetes Mellitus
[GWG]: Gestational Weight Gain
[LGA]: Large for Gestational Age
[WHO]: World Health Organization
[WSUTRH]: Wolaita Sodo University teaching and referral hospital

## ACKNOWLEDGEMENT

First of all, we would like to thank almighty God for His mercy and His assistance and we would like to thank Wolaita Sodo University, College of Health Sciences and Medicine. Finally, our gratitude goes to the study participants and data collectors.

